# Financing global health security: estimating the costs of pandemic preparedness in Global Fund eligible countries

**DOI:** 10.1101/2022.03.03.22271863

**Authors:** Stephanie Eaneff, Matthew R Boyce, Ellie Graeden, David W. Lowrance, Mackenzie Moore, Rebecca Katz

## Abstract

**Introduction:** The Global Fund to Fight AIDS, TB, and Malaria (the Global Fund) pivoted investments to support countries in their response to the COVID-19 pandemic. Recently, the Global Fund’s Board approved global pandemic preparedness and response as part of their new six-year strategy from 2023-2028.

**Methods:** Prior research estimated that US$124 billion is required, globally, to build sufficient country-level capacity for health security, with US$76 billion needed over an initial three-year period. Action-based cost estimates generated from that research were coded as directly, indirectly, or unrelated to systems strengthening efforts applicable to HIV, TB, and/or malaria.

**Results:** Of approximately US$76 billion needed for country level capacity-building over the next three-year allocation period, we estimate that US$66 billion is needed in Global Fund-eligible countries, and over one-third relates directly or indirectly (US$6 billion and US$21 billion, respectively) to health systems strengthening efforts applicable to HIV, TB, and/or malaria disease programs currently supported by the Global Fund. Among these investments, cost drivers include financing for surveillance and laboratory systems, to combat antimicrobial resistance, and for training, capacity-building, and ongoing support for the healthcare and public health workforce.

**Conclusion:** This work highlights a potential strategic role for the Global Fund to contribute to health security while remaining aligned with its core mission. It demonstrates the value of action-based costing estimates to inform strategic investment planning in pandemic preparedness.

**What is already known on this topic:**

- The costs, globally, to build country-level public health capacity to address these gaps over the next five years has been previously estimated as US$96-$204 billion, with an estimated US$63-131 billion in investments required over the next three years.
- Research conducted prior to the COVID-19 pandemic indicated that over one-third of Global Fund’s budgets in 10 case-study countries aligned with health security priorities articulated by the Joint External Evaluation, particularly in the areas of laboratory systems, antimicrobial resistance, and workforce development.

**What this study adds:**

- We estimate that over 85% of investments needed to build national capacities in health security, globally, over the next three years are in countries eligible for Global Fund support.
- Areas of investment opportunity aligned with the Global Fund’s core mandate include financing for surveillance and laboratory systems, combating antimicrobial resistance, and developing and supporting robust healthcare and public health workforces.

**How this study might affect research, practice or policy:**

- In aggregate, global-level data highlight areas of opportunity for the Global Fund to expand and further develop its support of global health security in areas aligned with its mandate and programmatic scope.
- Such investment opportunities have implications not only for existing budgeting and allocation processes, but also for implementation models, partners, programming, and governance structures, should these areas of potential expansion be prioritized.
- This work emphasizes a role for targeted, action-based cost estimation to identify gaps and to inform strategic investment decisions in global health.

## Introduction

The COVID-19 pandemic has demonstrated that the world was not well prepared to respond to an infectious disease threat of this magnitude. Countries across all socioeconomic and development categories have struggled to implement effective national responses. Substantial amounts of additional investment are required to support the development of country capacities to prevent, detect, and respond to both existing and emerging infectious disease threats. Prior research efforts have estimated that between US$96-$204 billion is required globally to advance national-level health security capacities with US$63-131 billion needed over a three-year period.[1-4] Given the substantial costs of ongoing COVID-19 response, recently estimated to be over US$12.5 trillion through 2024,[5] and an estimated 12·1 - 22·7 million excess deaths, globally, due to COVID as of January 2022,[6] the importance and potential return-on-investment of such upfront investment in capacity building is more evident than ever before.

The Global Fund to Fight AIDS, TB, and Malaria (the Global Fund) is a partnership between governments, private-sector organizations, civil society, and communities that supports health programs in over 120 countries and regions.[7] The Global Fund plays a major role in funding global health activities, and over the past two years has responded to the COVID-19 pandemic by pivoting many of its investments and personnel, procuring necessary diagnostics, personal protective equipment, and other essential health products, and supporting countries in their response to the virus through its COVID-19 Response Mechanism.[8] Recently, Global Fund’s Board approved pandemic preparedness and response as a pillar in their new six-year strategy from 2023-2028.[9]

Research conducted prior to the COVID-19 pandemic based on historical Global Fund budgeting patterns highlighted areas of opportunity for Global Fund investments to support global health security in domains including laboratory systems, workforce development, efforts to combat antimicrobial resistance, and the deployment of medical countermeasures, including key commodities and surge capacity.[10] Despite this previous work exploring the scope and focus of the Global Fund’s budget allocations before the pandemic, it remains challenging to prospectively assess what role the Global Fund might play in future efforts focused on building capacities for pandemic preparedness and response.

To examine this question, this research effort explores the relationship between investment requirements for health security and the Global Fund’s core mandates. More specifically, we examine the extent to which the Global Fund may be poised to help address national-level health security investment needs based on alignment with the Global Fund’s core mandate and current country eligibility requirements.

## Methods

### Cost estimation

Our team conducted prior research estimating that US$76 billion is required over three years to build health security capacity at the national and subnational levels, globally, based on anticipated investment needs for each of the 196 State Parties to the International Health Regulations (IHR).^1^ Briefly, public health capacity for each State Party was measured based on reported State Parties Self-Assessment Annual Reporting Tool (SPAR) assessment scores. Scores were mapped to benchmarks in the Joint External Evaluation (JEE)[11, 12] and analyzed alongside vaccination and workforce data from the WHO Global Health Observatory.[13, 14] Using the IHR Costing Tool,[15] costs were estimated for each State Party to achieve a score of “demonstrated capacity” (i.e., an assessment score of 4 out of a possible 5) on all indicators of the JEE, annually, over a five-year period. The results described hereafter correspond to the first three years of this investment period, selected to align with Global Fund’s three-year allocation period.

### Alignment of costed activities and Global Fund mandate

Based on prior estimates of investment requirements, an in-depth analysis was completed to identify which of over 700 activities contributed directly or indirectly to health systems strengthening efforts in support of HIV, TB, and/or malaria (HTM) disease programs. Existing activities were reviewed by a minimum of two members of the research team and tagged based on (1) whether the investment was required in a Global Fund-eligible country and (2) whether the investment contributed directly, indirectly, or was unrelated to HTM efforts.

For the purposes of this study, countries were considered eligible for Global Fund support if they were “eligible” or “in transition” to receive funding for one or more disease components (i.e., HTM) in 2021.[16] Direct contributions were defined as interventions, activities, or resources critical to the delivery of quality HTM services,[17] including support for skilled health and public health workers whose work is significantly focused on these disease areas (e.g., personnel trained to identify and manage infections caused by AMR resistant pathogens); indirect contributions were defined as interventions, activities, or resources focused primarily on non-HTM disease areas, but that could be pivoted to use directly toward HTM efforts during times of need (e.g., diagnostics for COVID-19 that could be repurposed for TB); unrelated investments were considered to be interventions, activities, or resources that do not directly or indirectly contribute to HTM efforts, including interventions or materials that support non-HTM disease areas and/or that could not be rapidly pivoted directly toward HTM-related efforts. Of note, in the case of investments identified as “directly” or “indirectly” related to HTM efforts, the assumption was made that resources could, and would, be able to be pivoted during times of need to support multiple disease areas, including HTM. Such investments included skilled healthcare and public health workforce support, general consumable laboratory materials and laboratory training, transportation resources, and personal protective equipment that could be shared to support cross-cutting efforts across disease verticals during times of need. Table 1, below, provides select examples of interventions tagged as directly, indirectly, and unrelated to HTM efforts.

**Table 1.** Selected examples of activities identified as directly, indirectly, and unrelated to HIV, TB, and/or malaria (HTM) efforts.

| Relationship to HTM efforts | Type of activity |
| --- | --- |
| Directly related to HTM efforts | Skilled healthcare and public health trained in surveillance and infection prevention and control |
|  | Select rapid diagnostic tests and point of care diagnostics |
|  | Support for national-level supply chain capacity, including warehouse space to send, receive, and maintain an inventory of medical countermeasures |
| Indirectly related to HTM efforts | Laboratory equipment and consumable materials associated with virus culture, serology, and PCR capabilities, including genetic analyzers and real-time PCR systems |
|  | Support for national-level risk communication specialists and broadcast time for proactive health communication |
|  | Stipends and supplies for annual cohorts of full-time field epidemiology training programs (FETP) trainees |
| Unrelated interventions | Personnel and supplies to respond to chemical and/or radiological emergencies |
|  | Physical infrastructure for Emergency Operations Center (EOC) |

### Aggregation and global analysis

Individual action-based costs were aggregated to identify trends, cost drivers, and potential areas of opportunity for the Global Fund. Costs were summarized globally, consistent with the intended use of cost estimations to provide high-level global estimates, as opposed to detailed country-level action plans, which are best developed and informed by local public health expertise. Since analyses did not estimate costs beyond a score of “demonstrated capacity,”, IHR member states already reporting such scores do not have any additional costs for these respective capacities. Given the order-of-magnitude nature of cost estimates, reported costs are rounded to the nearest billion and/or the nearest percentage point; costs under US$1 billion are rounded to the nearest US$100 million. This may introduce minor inconsistencies in reported numbers due to rounding approximation. All costs were reported in 2021 US$.

### Patient and Public Involvement

Given the nature of this cost-based analysis, patients or the public were not involved in the design, or conduct, or reporting of the current research. The IHR costing tool, which methodologically underlies the current work, was informed by in-depth case studies conducted in countries spanning multiple regions. Based on user feedback from a global user base, the tool’s methods have been iteratively updated to reflect changing assumptions, understandings, and developments in preparedness metrics.

## Results

In countries eligible to receive Global Fund support for HTM, an estimated US$66 billion in investment is required to develop “demonstrated capacity” in global health security over an initial three years of funding. This investment requirement in over 120 eligible countries comprises over 85% of the US$76 billion required, globally, to fund national-level capacity in the same time period. Of global cost needs (US$76 billion), approximately US$27 billion (36%) relates either directly or indirectly (US$6 billion and US$21 billion, respectively) to systems strengthening in support of HTM efforts in Global Fund eligible countries (Figure 1).

**Figure 1.**
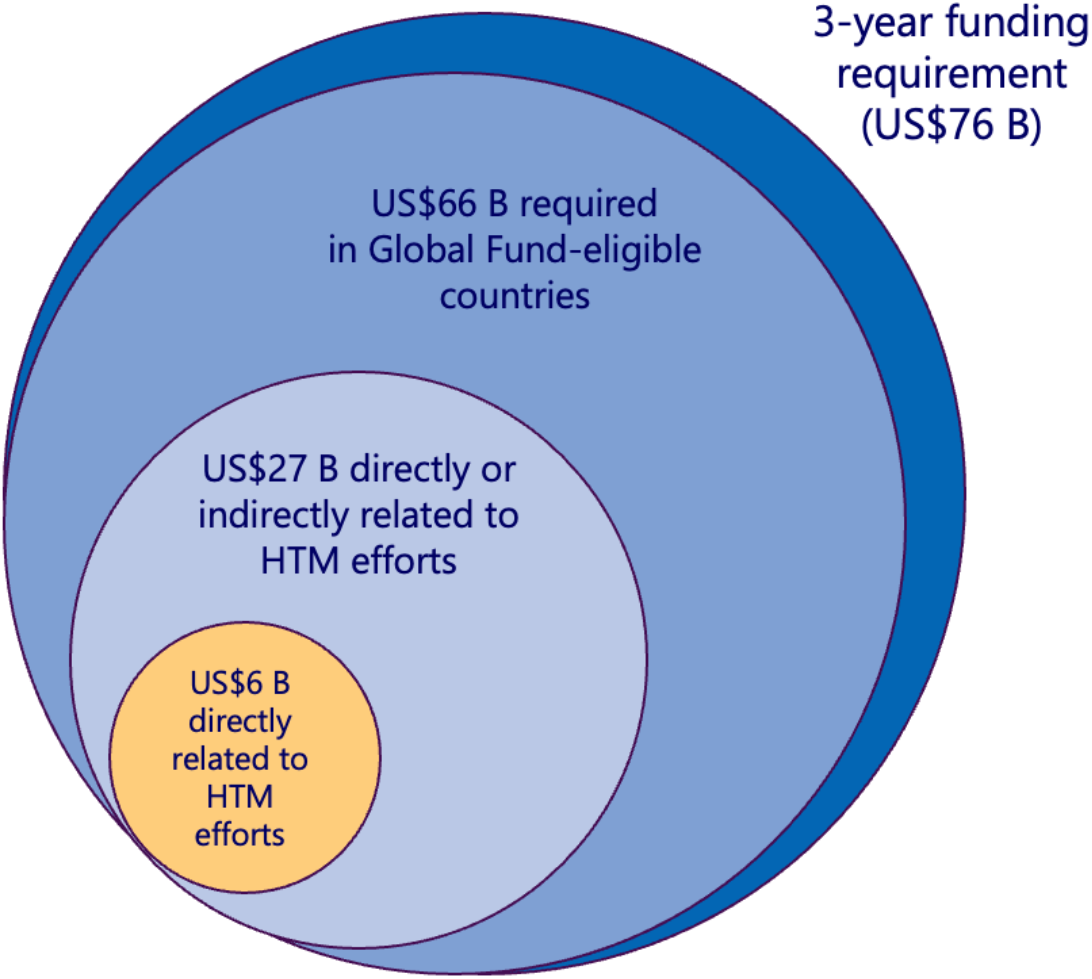
Proportion of global costs over a three-year period that occur in Global Fund eligible countries and that are directly and/or indirectly related to HIV, TB, and/or malaria (HTM) efforts.

Approximately US$2 billion is required annually for costs directly related to HTM efforts over the first three years of financing (Table 2). For costs indirectly related to HTM efforts, approximately US$6 billion is required in the first year, with an additional US$7 billion required in the second year, and an additional US$8 billion required in the third. These costs gradually increase over time primarily because of costs related to scaling-up personnel capacities.

**Table 2.**
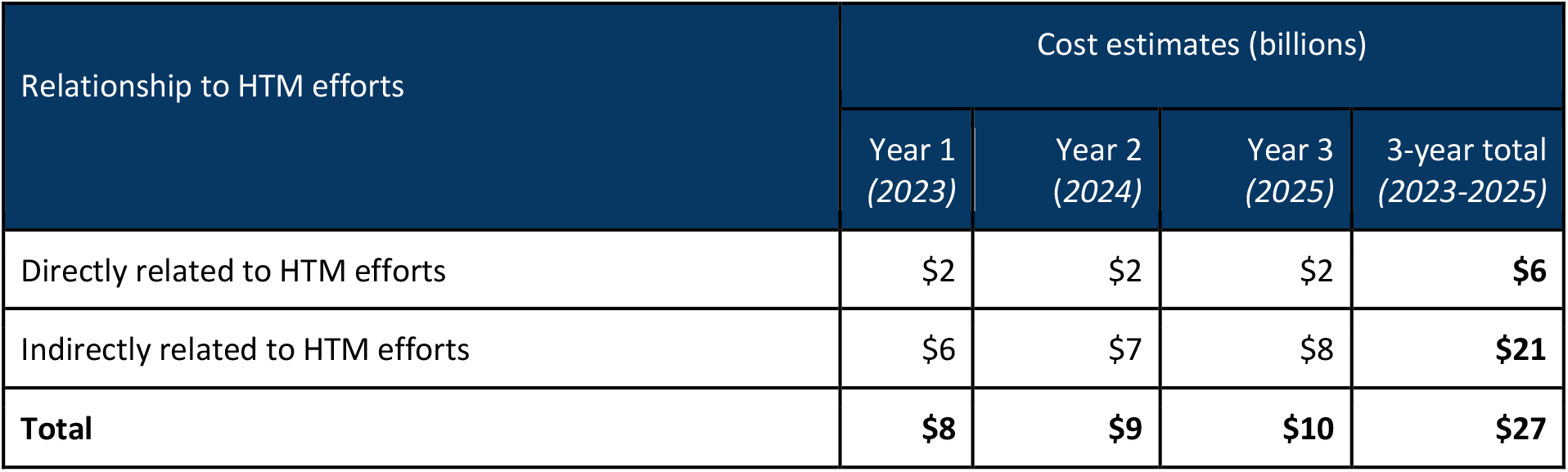
Estimated costs directly or indirectly related to HTM efforts in Global Fund eligible countries over a three-year funding cycle, assuming all funding initiated in 2023. All costs estimated in 2021 US$.

Clear cost drivers eligible for future Global Fund support include capacity building for surveillance and laboratory systems; such systems would also support broader health security objectives given that they are designed and implemented in a way that allows for them to be used across disease areas. Similarly, efforts to combat antimicrobial resistance are also eligible for substantial amounts of support and have the potential to broadly strengthen health systems if implemented cross-functionally. Within each of these areas, costs for the training, development, and sustained support of personnel are the greatest contributor to total costs. These reflect the requirements for developing and maintaining a robust workforce of skilled public health, healthcare, and community health workers. Table 3, below, identifies a select list of areas, aligned with the JEE core capacities, with significant investment opportunities that are directly or indirectly related to the Global Fund’s mandate and programmatic scope. Of note, while surveillance, laboratory, and antimicrobial resistance capacities comprise a significant proportion of the global costs identified as directly or indirectly relevant to HTM efforts, cross-functional investment requirements identified (i.e., those across core capacities) include risk communication, emergency response operations, and support for national legislation, policy, and financing.

**Table 3.** Select investments that are directly or indirectly related to HIV, TB, and/or malaria (HTM) efforts, by core capacity of the JEE. All costs estimated in 2021 US$; results reported rounded to the nearest billion or million, depending on the order of magnitude. Selected activities are intended as illustrative examples but do not cover the full costs for each core capacity; each activity is costed as part of a single core capacity and individual activities and costs are not duplicated across core capacities. Of note, these costs represent overlap between investments related to HTM efforts and need as assessed for progress against the specified JEE indicators; as such, they do not represent the total magnitude of global need in each area.

| JEE core capacity | 3-year costs<br>(directly & indirectly<br>related to HTM<br>efforts) | Selected examples of costed activities<br>(directly & indirectly related to HTM efforts) |
| --- | --- | --- |
| Real-time surveillance | US\$13 billion | *Training, capacity building, and ongoing support for skilled healthcare and public health workforce to enable both indicator and event-based surveillance; Development and maintenance of electronic disease surveillance data platforms* |
| Antimicrobial Resistance | US\$8 billion | *Resources to support and enable infection prevention and control (IPC) in healthcare facilities, including decontamination kits, airborne infection isolation rooms, and hand hygiene kits; outbreak investigation kits; training, development, and ongoing support for skilled healthcare and public health workforce* |
| National Laboratory System | US\$3 billion | *Durable & consumable laboratory materials that support efforts aligned with multiple disease areas (e.g., virus culture, serology, and PCR capability, microscopy and bacterial culture capability); select rapid diagnostic tests* |
| Workforce Development | US\$1 billion | *Expand national field epidemiology training (FETP) capability and provide programmatic support and essential supplies for training; develop, maintain, and evaluate national workforce strategy* |
| Risk communication | US\$800 million | *Risk communication planning; development and implementation of community level engagement strategies; fees for communication via newspaper, radio, and television; infrastructure for dynamic listening and rumor management* |
| Medical Countermeasures and Personnel Deployment | US\$400 million | *Storage facilities to receive, store, and distribute durable and consumable materials during times of acute need, including support for the cold chain; Development of plans and exercises to develop national and subnational capacity for medical countermeasure and surge personnel deployment* |
| Emergency Response Operations | <US\$100 million | *Development of incident management plans, including data collection, reporting, and briefing; Functional exercises of emergency operations center (EOC) activation; coordination with public health communication functions* |
| National legislation, policy, and financing | <US\$100 million | *Assess implementation of IHR-relevant legislation and policies, including review and revision of cross-border agreements or protocols* |

Investment requirements are impacted by both existing preparedness capacities and by the costs required to strengthen capacities. Costs directly or indirectly related to HTM efforts accumulate in domains where current global capacity is lowest, in domains that require high-cost investments, and in domains addressing activities and resources that are directly critical to the delivery of HTM services. The indicators that drive costs most substantially, including antimicrobial resistance, are those with low global capacity and high-cost investment requirements. For instance, 80% of reporting countries earned an average score below “demonstrated capacity” for combating antimicrobial resistance on the JEE.[18] Efforts to combat antimicrobial resistance require expensive investment in personnel and in resources, including healthcare facility-level investments in infection prevention and control (e.g., hand washing equipment, decontamination kits, and airborne isolation rooms); these investments relate directly to the Global Fund’s mandate and programmatic scope, and are essential for supporting the delivery of quality HTM services.

## Conclusion

Taken together, these findings suggest that the Global Fund is well positioned to take a leadership role in financing certain areas of global health security and pandemic preparedness and response, given additional consideration for appropriate implementation models, partners, programming, and governance structures. We estimate that over 85% of national-level financing requirements, globally, over the first three years occur in countries eligible for Global Fund support, and that over one third of those costs directly or indirectly relate to HTM efforts. Estimates highlight significant areas of opportunity for the Global Fund’s role in global health security financing, particularly in low-and-middle income countries eligible for Global Fund support. These substantial investment opportunities come hand in hand with considerations for implementation. Translating budget alignment to implementation impact in health security will require adjustments and the adaptation of existing practices – new and expanded governance structures, additional technical expertise and coordination efforts, and a systems approach to implementation across vertical disease verticals.

Surveillance, laboratory, and antimicrobial resistance functions comprise a significant proportion of health security investment requirements that were identified as directly or indirectly relevant to HTM efforts. However, they are certainly not the only areas where critical capacity strengthening resources are needed. Additional areas of opportunity include support for human resources and workforce development, risk communication and community engagement, and supply chain management, including support for the cold chain. Notably, these investments are required to support multiple disease areas and efforts beyond HTM. Cross-functional implementation that simultaneously supports the needs of multiple disease verticals has the potential to bolster efforts to combat HTM while also helping promote a state of readiness to respond to other infectious disease threats and emergencies as they arise.

This report has several important limitations. The WHO has recently revised the SPAR/JEE framework that was the basis of the activity-based costing. However, the updated versions of these tools have not yet been officially released at the time of this analysis. The original, costed version of the JEE did not fully reflect some components of pandemic preparedness, including investment in water, sanitation and hygiene (WASH), robust infection prevention and control (IPC) activities, and support for community health workers, which are likely to be important cost drivers in future operational plans. Additionally, it is important to note that the cost estimates referenced in this analysis represent the overlap between the Global Fund’s programmatic scope and global need as assessed by the JEE; these estimates are not representative of the magnitude of either overall cost estimate, individually. Furthermore, increased health financing for pandemic preparedness via the Global Fund would hold important implications for country governance and planning; implementation arrangements; monitoring and accountability; and partnerships, which are beyond the immediate scope of this analysis.

In sum, these results underscore the potential of country-level, action-based costing analyses to inform medium and long-term strategic investment planning in global health security. Specific, national and subnational investment requirements are best informed by local expertise and context-specific knowledge of imminent risks, response performance gaps, and community needs. However, in aggregate, global-level data highlight clear areas of opportunity for the Global Fund to expand its support for health security and pandemic preparedness and response with a transparent, representative, and country-driven approach.

## Data Availability

Aggregate data that underlie the results reported in this article will be made available to those conducting research or supporting operations that are consistent with the intended use of these data for global cost estimation. Proposals and data inquiries should be directed to the corresponding author.

https://ghscosting.org/

## Author Contributorship

SE, MB, EG, MM, and RK contributed to study conceptualization and design. SE, MB, MM, DL, and RK made significant contributions toward data analysis, and all authors contributed towards the interpretation of results. SE created the figures and drafted the manuscript that was subsequently reviewed and revised by all authors. All authors have read and approved of the final version of the manuscript.

## Acknowledgements

The authors wish to thank Richard Grahn (Global Fund), Johannes Hunger (Global Fund), and Mehran Hossini (Global Fund) for their collaboration and feedback on the interpretation of data, and Hailey Robertson (Talus Analytics) for assistance with data collection and tagging.

